# On the changing role of individuals in different age groups in propagating the Omicron epidemic waves in France

**DOI:** 10.1101/2022.12.22.22283867

**Authors:** Edward Goldstein

**Affiliations:** Massachusetts Eye and Ear, Harvard Medical School, Boston, MA 02114, USA

## Abstract

**Objectives:** In late December 2022, rates of mortality in France (over 2,500 daily deaths) have reached levels not seen since April 2020, with the most pronounced increase in mortality recorded in nursing homes. Epidemics of Omicron and influenza have both contributed to those high mortality levels in late 2022. The roles of different age groups in propagating Omicron epidemics in the whole community require a better characterization, particularly given that in France, vaccination coverage for the 2nd booster for COVID-19 is limited and largely restricted to persons aged over 60y.

**Patients and Methods:** We examined the role of individuals in different age groups in propagating different waves of Omicron epidemics in France between March 1--Dec. 30, 2022 using previously developed methodology based on the relative risk (RR) statistic that measures the change in the group’s proportion among all cases before vs. after the peak of an epidemic wave. Higher value of the RR statistic for a given age group suggests a disproportionate depletion of susceptible individuals in that age group during the epidemic*’*s ascent (due to increased contact rates and/or susceptibility to infection).

**Results:** For the Spring wave (March 14 - May 15), the highest RR estimate belonged to children aged 10-19y (RR=1.92 (95% CI (1.18,3.12)), followed by adults aged 40-49y (RR=1.45 (1.09,1.93)) and children aged 0-9y (RR=1.31 (0.98,1.74)). For the Summer wave (June 27 – Aug. 21), the highest RR estimate belong to children aged 0-9y (RR=1.61 (1.13,2.30)) followed by children aged 10-19y (RR=1.59 (0.77,3.26)) and adults aged 20-29y (RR=1.42 (0.91,2.23)). For the Autumn wave (Sep. 18 – Nov. 12), the highest RR estimate belonged to children aged 10-19y (RR=1.65 (0.72,3.75)), followed by adults aged 30-39y (RR=1.39 (0.83,2.33)) and 20-29y (RR=1.21 (0.66,2.23)). For the Autumn-Winter Wave (Nov. 23 – Dec. 30), the highest RR estimate belonged to persons aged 30-39y (RR=1.43 (0.79,2.57)), followed by persons aged 80-89y (RR= 1.17 (0.99,1.4)) and persons aged 40-49y (RR= 1.15 (0.73,1.82))).

**Discussion/Conclusions:** Increasing booster vaccination coverage for all adults, as possibly for children should help mitigate future Omicron epidemics. The estimate for the RR statistic in persons aged 80-89y for the Autumn-Winter Omicron wave suggests that additional efforts should be considered for preventing the spread of Omicron infection in elderly persons, including in Long-Term Care facilities.

## Introduction

During December 2020, high levels of circulation of Omicron and influenza took place in France. The peak of the Omicron epidemic took place around weeks 49-50 (as suggested by data on confirmed cases [1]), whereas the peak of the influenza epidemic took place around week 52 [2]. High rates of all-cause mortality, not seen since April 2020, were recorded in France during the 2^nd^ half of December 2022, with the 1^st^ peak of 2,531 daily deaths recorded on Dec. 19 and the 2^nd^ peak of 2,522 all-cause daily death recorded on Dec. 26 [3]. Moreover, data on the place of death suggests that the greatest relative increases in mortality in the 2^nd^ half of December 2022 in France took place in long-term care facilities for older individuals [4]. Those increases in mortality, particularly during the periods of the Omicron epidemic waves, suggest the need to better understand the role of different population groups in propagating Omicron epidemics in the community with the aim of informing booster vaccination policies and other mitigation measures in different population groups.

With the emergence of the Omicron variant of SARS-CoV-2, the risk of complications, including hospital admission and death in adults became lower compared to the Delta variant, though those relative risks vary with age, with the relative risk for severe outcomes, including death for Omicron vs. Delta being greatest for the oldest adults [5,6]. In addition to changes in severity, emergence of the Omicron variant brought about changes in the tole of individuals in different age groups in the transmission of SARS-CoV-2 infection in the community. During the circulation of earlier SARS-CoV-2 variants, the leading role in the acquisition and spread of infection process generally belonged to younger adults (aged 18-35y) and older adolescents [7-10] – in particular, see the temporal data on the prevalence of SARS-CoV-2 infection in different age groups in England in [10]. The appearance of the Omicron variant resulted in a greater role of children in the transmission process, at least during the early stages of the Omicron epidemic [11]. For example, findings from REACT-1 study in England for samples tested between January 5-20, 2022 saw the greatest prevalence of infection in children aged 5-11y [12] (which is different from what was observed for the earlier SARS-CoV-2 variants [10]). The role of different age groups during subsequent waves of Omicron epidemics needs a better characterization (e.g. see the Coronavirus Infection Surveys in the UK [13]). Such information could inform efforts of booster vaccination, as well as other mitigation efforts aimed at limiting the spread of Omicron infection in the community. We note that there is evidence that vaccination reduces both the risk of acquisition of Omicron infections [14], as well as the risk of onward transmission of infection [15]. Additionally, vaccination coverage for the 2^nd^ booster for COVID-19 in France is limited and largely restricted to persons aged over 60y [16].

In [17-19] we developed a method for assessing the relative role (per average individual) in different subpopulations in the spread of infection during epidemics of infectious diseases. The idea of that method is that subpopulations that play a disproportionate role during the outbreak*’*s ascent due to increased susceptibility to infection and/or contact rates can be related to the relative risk (RR) statistic that evaluates the change in the subpopulation*’*s proportion among all cases in the population before vs. after the epidemic*’*s peak (see Methods). Moreover, we used simulations in the context of influenza epidemics ([17]) to show a relation between a higher value for the RR statistic in a given age group and a higher impact of vaccinating an individual in that age group on reducing the epidemic*’*s growth rate. This method was also used in the context of SARS-CoV-2 epidemics in 2020 and 2021 to show the prominent relative role of younger adults and older adolescents in the transmission process [8,9]. In this paper we use the RR statistic to examine the role of individuals in different age groups during four waves of the Omicron epidemic in France between March 1, 2022 and Dec. 30, 2022. The goal is to characterize the role of individuals in different age groups in the spread of infection during the different waves of the Omicron epidemic beyond the initial (2022 winter) wave to inform vaccination efforts, and other efforts aiming at mitigation of Omicron transmission in the community.

## Methods

### Data

Data on the rates of ICU admissions for COVID-19 in different age groups in France are available in [1,20]. Data on the age structure of the population in France in 2022 are available in [21]. Those data were combined to evaluate the numbers of ICU admissions during the previous week for each day between March 1, 2022 and Dec. 30, 2022.

### Statistical Inference

Based on the data for ICU admissions in all age groups in France (Figure 1), we delineated four epidemic waves between March 1, 2022 and Dec. 17, 2022: The Spring wave (March 14 - May 15, peak day for ICU admissions being April 14), the Summer wave (June 27 – Aug. 21, peak day for ICU admissions being July 21), the Autumn wave (Sep. 18 – Nov. 12, peak day for ICU admissions being Oct. 19), and the Autumn-Winter wave (Nov. 23 – Dec. 30, peak day for ICU admission being Dec. 17). For each epidemic wave, we excluded the 7-day period around the peak day for ICU admissions, and defined the before-the-peak period for that epidemic wave as the period from the start of the epidemic wave to the last day before the 7-day window around the peak day for ICU admissions -- thus, for the Spring epidemic wave between March 14 - May 15, with the peak day for ICU admissions being April 14, the before-the-peak period of the wave is March 14 – April 10, etc. For each epidemic wave except for the Autumn-Winter wave, we defined the after-the-peak period of the epidemic wave as the period starting from the first day after the 7-day window around the peak of ICU admissions to the end of the epidemic wave --- thus, for the Spring epidemic wave between March 14 - May 15, with the peak day for ICU admissions being April 14, the after-the-peak period of the wave is April 18 – May 15, etc. For the Autumn-Winter wave, patterns of transmission of infection have changed as schools closed for the winter break, and we defined the after-the-peak period for ICU admissions as the period between Dec. 17—Dec. 30.

**Figure 1:**
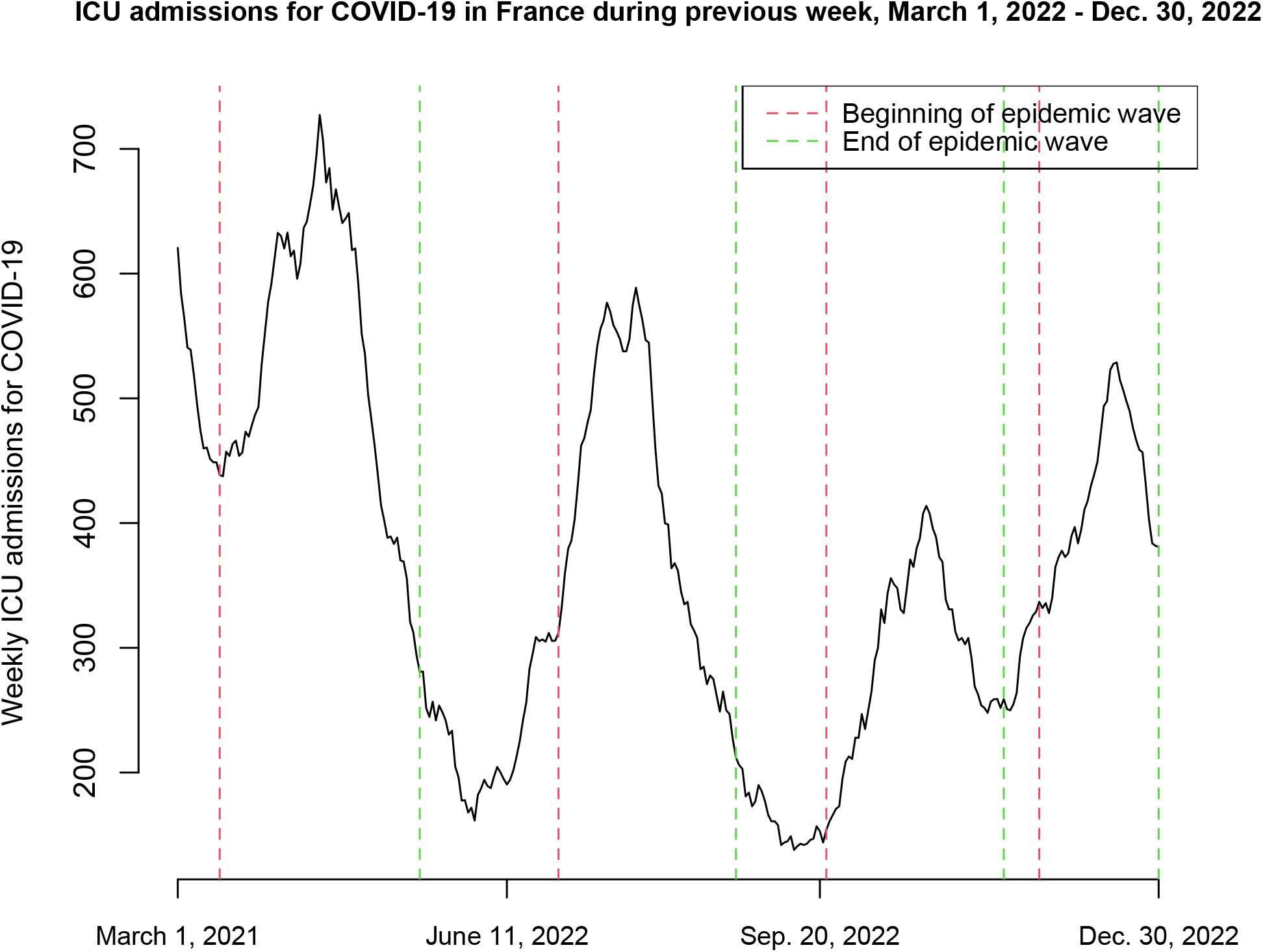
ICU admissions for COVID-19 in France during the previous week, March 1, 2022 - Dec. 30, 2022, and the four epidemic waves (March 14 - May 15, June 27 – Aug. 21, Sep. 18 – Nov. 12, Nov. 23 - Dec. 30).

We considered 10 age groups in our analyses: 0-9y, 10-19y, 20-29y, 30-39y, 40-49y, 50-59y, 60-69y, 70-79y, 80-89y, 90+y. For each epidemic wave, and each age group *g*, let *B*(*g*) be the number of ICU admissions for COVID-19 in persons in age group *g* during the before-the-peak-period, and let *A*(*g*) be the number of ICU admissions for COVID-19 in persons in age group *g* during the after-the-peak-period. The proportion of cases belonging to the age group *g* among all ICU admissions for COVID-19 during the before-the-peak period is therefore

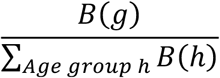

The relative risk (RR) statistic measures the change in the group*’*s proportion among all cases admitted to ICU before vs. after the peak of the epidemic wave. The point estimate for the relative risk *RR*(*g*) in an age group *g* is:

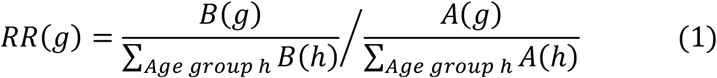

We note that the relative magnitudes of the RR statistic in different age groups do not depend on the proportion of cases of Omicron infection in different age groups that end up in an ICU. This is because for each age group,

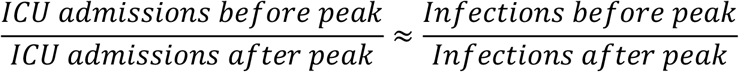

The reason for using data on ICU admissions for COVID-19 rather than data on detected cases of COVID-19 is that testing for SARS-CoV-2 infection in symptomatic cases is much more consistent in the ICU setting compared to ambulatory/community testing of cases.

Higher value of the RR statistic for a given age group suggests a disproportionate depletion of susceptible individuals in that age group during the epidemic*’*s ascent (due to increased contact rates and/or susceptibility to infection), resulting in the decline in the share of that age group among all cases during the epidemic*’*s descent. We assume that the numbers of reported cases are sufficiently high so that the logarithm ln(RR(g)) of the relative risk RR in the age group g is approximately normally distributed [22]. Under this approximation, the 95% confidence interval for RR (g) is exp(ln(RR(g)) ± 1.96 · SE(g)), where ln(RR(g)) is estimated via eq. 1, and the standard error SE(g) is ([22]):

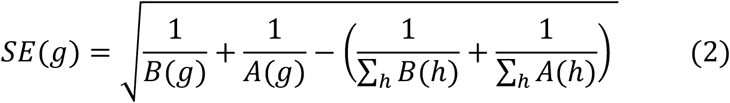

## Results

Figure 1 plots the numbers of ICU admissions for COVID-19 in France during the previous week between March 1, 2022 - Dec. 30, 2022, including the four study periods used in our analysis (March 14 - May 15, June 27 – Aug. 21, Sep. 18 – Nov. 12, Nov. 23 - Dec. 30). Plots of the rates of ICU admissions in different age groups are contained in [1].

Table 1 gives the estimates of the relative risk (RR) statistic in different age groups during the Four epidemic waves considered in our study. For the Spring wave (March 14 - May 15), the highest RR estimate belonged to children aged 10-19y (RR=1.92 (95% CI (1.18,3.12)), followed by adults aged 40-49y (RR=1.45 (1.09,1.93)) and children aged 0-9y (RR=1.31 (0.98,1.74)). For the Summer wave (June 27 – Aug. 21), the highest RR estimate belong to children aged 0-9y (RR=1.61 (1.13,2.30)) followed by children aged 10-19y (RR=1.59 (0.77,3.26)) and adults aged 20-29y (RR=1.42 (0.91,2.23)). For the Autumn wave (Sep. 18 – Nov. 12), the highest RR estimate belonged to children aged 10-19y (RR=1.65 (0.72,3.75)), followed by adults aged 30-39y (RR=1.39 (0.83,2.33)) and 20-29y (RR=1.21 (0.66,2.23)). For the Autumn-Winter Wave (Nov. 23 – Dec. 30), the highest RR estimate belonged to persons aged 30-39y (RR=1.43 (0.79,2.57)), followed by persons aged 80-89y (RR= 1.17 (0.99,1.4)) and persons aged 40-49y (RR= 1.15 (0.73,1.82))).

**Table 1:** Estimates for the relative risk RR (eq. 1) for data on ICU admissions in different age groups during four Omicron waves in France in 2022: March 14 - May 15, June 27 – Aug. 21, Sep. 18 – Nov. 12, Nov. 23 - Dec. 30, 2022

| Age group | Spring wave<br>(Mar. 14 – May 15) | Summer Wave<br>(June 27 – Aug. 21) | Autumn Wave<br>(Sep. 18 – Nov. 12) | Autumn-Winter Wave<br>(Nov. 23 – Dec. 30) |
| --- | --- | --- | --- | --- |
| 0-9y | 1.31 (0.98,1.74) | 1.61 (1.12,2.3) | 0.79 (0.53,1.19) | 0.98 (0.67,1.43) |
| 10-19y | 1.92 (1.18,3.12) | 1.59 (0.77,3.26) | 1.65 (0.72,3.75) | 0.67 (0.28,1.61) |
| 20-29y | 0.93 (0.62,1.39) | 1.42 (0.91,2.23) | 1.21 (0.66,2.23) | 0.77 (0.38,1.57) |
| 30-39y | 0.96 (0.68,1.36) | 0.89 (0.62,1.29) | 1.39 (0.83,2.33) | 1.43 (0.79,2.57) |
| 40-49y | 1.45 (1.09,1.93) | 1.03 (0.75,1.41) | 0.92 (0.61,1.4) | 1.15 (0.73,1.82) |
| 50-59y | 1.16 (0.97,1.39) | 0.89 (0.72,1.09) | 1 (0.75,1.32) | 0.94 (0.73,1.22) |
| 60-69y | 0.86 (0.76,0.97) | 0.9 (0.77,1.05) | 1.05 (0.87,1.26) | 0.95 (0.79,1.14) |
| 70-79y | 0.93 (0.85,1.02) | 1.02 (0.91,1.15) | 0.87 (0.76,1) | 0.93 (0.82,1.05) |
| 80-89y | 0.97 (0.84,1.11) | 1 (0.85,1.17) | 1.19 (1,1.42) | 1.17 (0.99,1.4) |
| 90+y | 1.10 (0.81,1.5) | 0.89 (0.63,1.27) | 0.76 (0.5,1.16) | 1.09 (0.72,1.65) |

## Discussion

High rates of all-cause mortality, not seen since April 2020, were recorded in France during the 2^nd^ half of December 2022 [3], with some of the excess mortality being associated with the Omicron epidemic in France during that period [1] (with an influenza epidemic peaking about 2-3 weeks after the Omicron epidemic also making a contribution to the mortality burden [2]).

Those elevated mortality levels related to the Omicron epidemic suggest the need to better understand the role of different population groups in propagating Omicron epidemics in the community with the aim of informing booster vaccination policies and other mitigation measures in different population groups. In this paper, we applied the previously developed methodology [17-19,7,8] to study the relative role of individuals in different age groups during the Spring, Summer, Autumn and Autumn-Winter waves of the 2022 Omicron epidemic in France. We found that children aged 10-19y played the greatest relative role in propagating Spring and the first Autumn waves of the Omicron epidemics, while for the Summer wave (when schools were closed), the estimate of the RR statistic in children aged 10-19y was slightly lower compared to those aged 0-9y. We note that several studies have documented large SARS-CoV-2 outbreaks in the school setting under limited mitigation [23-25], which is generally pertinent to the Omicron period compared to earlier periods when different mitigation (including social distancing) measures in schools were put in place. Following children aged 10-19y and 0-9y, the most prominent role in propagating the Spring, Summer, and Autumn waves of the 2022 Omicron epidemic belonged to adults aged 20-29y and adults aged 30-49y. For the Autumn-Winter Omicron epidemic wave in France, the greatest depletion of susceptible individuals during the ascent period of the epidemic was in persons aged 30-39y, followed by persons aged 80-89y and 40-49y, with the change compared to the previous epidemic waves presumably having to do with the built-up of immunity to Omicron in children and younger adults during those previous epidemic waves. Overall, our results suggest that increase in booster vaccination coverage in all adults, and possibly in children, should help limit the spread of infection during future Omicron epidemics in the community. We note that there is evidence that vaccination, including the use of boosters reduces the risk of acquisition of Omicron infections [14], as well as the likelihood of the onward spread of infection [15]. At the same time, vaccination coverage for the 2^nd^ booster in France in still limited and largely restricted to persons aged over 60y [1,16]. Additionally, active depletion of susceptible individuals in persons aged 80-89y during the ascent period of the Autumn-Winter wave of the Omicron epidemic in France in 2022 (found using the RR statistic in this paper), together with the increase in mortality in long-term care facilities for older individuals in France during that period (greater than increases in mortality in other settings in France at that time [4]) suggests that further efforts should be undertaken for limiting the spread of infection in the elderly, including in Long-Term Care Facilities.

Our results have some limitations. Inconsistency in testing/detection of Omicron infections would affect the estimates of the relative risk (RR) statistic. We used data on ICU admissions for COVID-19 rather than hospitalizations for COVID-19 or detected cases of COVID-19 in the community since under-detection of SARS-CoV-2 infection is much less pronounced in symptomatic ICU admissions compared to hospitalizations and ambulatory/community testing of cases. There is uncertainty regarding the relation between the RR statistic and the role that individuals in different age groups play in propagating epidemics. We used simulations in the context of influenza epidemics ([17]) to show a relation between a higher value for the RR statistic in a given age group and a higher impact of vaccinating an individual in that age group on reducing the epidemic*’*s initial growth rate. For adults aged over 80y, the relatively high estimate of the RR statistic during the Autumn-Winter wave of the Omicron epidemic in France likely suggests active spread of Omicron infection in elderly individuals, possibly in part in Long-Term Care facilities.

## Conclusions

As time progressed, the leading relative roles (per individual) in propagating Omicron epidemic waves in France shifted from children (particularly children aged 10-19y) and younger adults to other age groups of adults, particularly persons aged 30-49y during the Autumn-Winter Omicron wave in 2022. Increasing booster vaccination coverage for COVID-19 for all adults [1,16], and possibly for children should help mitigate the spread of Omicron infection in the community. Additionally, further efforts should be considered for preventing the spread of Omicron infection in elderly persons, including in Long-Term Care facilities.

## Data Availability

This study is based on aggregate, de-identified, publicly available data that can be accessed through refs. 1,20,21

https://www.santepubliquefrance.fr/dossiers/coronavirus-covid-19/coronavirus-chiffres-cles-et-evolution-de-la-covid-19-en-france-et-dans-le-monde

https://www.data.gouv.fr/fr/datasets/donnees-hospitalieres-relatives-a-lepidemie-de-covid-19/

https://www.insee.fr/fr/statistiques/2381472#graphique-Donnes

